# CarD-T: Interpreting Carcinomic Lexicon via Transformers

**DOI:** 10.1101/2024.08.13.24311948

**Authors:** Jamey O’Neill, Gudur Ashrith Reddy, Nermeeta Dhillon, Osika Tripathi, Ludmil Alexandrov, Parag Katira

## Abstract

The identification and classification of carcinogens is critical in cancer epidemiology, necessitating updated methodologies to manage the burgeoning biomedical literature. Current systems, like those run by the International Agency for Research on Cancer (IARC) and the National Toxicology Program (NTP), face challenges due to manual vetting and disparities in carcinogen classification spurred by the volume of emerging data. To address these issues, we introduced the Carcinogen Detection via Transformers (CarD-T) framework, a text analytics approach that combines transformer-based machine learning with probabilistic statistical analysis to efficiently nominate carcinogens from scientific texts. CarD-T uses Named Entity Recognition (NER) trained on PubMed abstracts featuring known carcinogens from IARC groups and includes a context classifier to enhance accuracy and manage computational demands. Using this method, journal publication data indexed with carcinogenicity & carcinogenesis Medical Subject Headings (MeSH) terms from the last 25 years was analyzed, identifying potential carcinogens. Training CarD-T on 60% of established carcinogens (Group 1 and 2A carcinogens, IARC designation), CarD-T correctly to identifies all of the remaining Group 1 and 2A designated carcinogens from the analyzed text. In addition, CarD-T nominates roughly 1500 more entities as potential carcinogens that have at least two publications citing evidence of carcinogenicity. Comparative assessment of CarD-T against GPT-4 model reveals a high recall (0.857 vs 0.705) and F1 score (0.875 vs 0.792), and comparable precision (0.894 vs 0.903). Additionally, CarD-T highlights 554 entities that show disputing evidence for carcinogenicity. These are further analyzed using Bayesian temporal Probabilistic Carcinogenic Denomination (PCarD) to provide probabilistic evaluations of their carcinogenic status based on evolving evidence. Our findings underscore that the CarD-T framework is not only robust and effective in identifying and nominating potential carcinogens within vast biomedical literature but also efficient on consumer GPUs. This integration of advanced NLP capabilities with vital epidemiological analysis significantly enhances the agility of public health responses to carcinogen identification, thereby setting a new benchmark for automated, scalable toxicological investigations.

## Introduction

The ongoing endeavor to discern and classify carcinogenic agents constitutes a critical facet of public health, with implications for risk assessment and the formulation of preventive measures^1^. Foundational repositories maintained by groups such as the United States National Toxicology Program^2^ (NTP), and the International Agency for Research on Cancer (IARC)^3,4^ are pivotal, offering well-documented research behind the classifications that encapsulate the carcinogenic potential of agents, grounded in rigorous scientific scrutiny. Yet, the current exponential growth of biomedical literature places an immense strain on authorities tasked with classifying and regulating carcinogens, such as the Occupational Safety and Health Administration (OSHA)^5^, Environmental Protection Agency (EPA)^6^, and the Europeans Chemicals Agency (ECHA)^7^, struggling to keep pace with the rapid dissemination of new findings^8–10^.

Presently, across the repositories published by IARC, NTP, EPA, ECHA and OSHA, a total of 480 unique causal entities or hazards for carcinogenesis are identified. However, a lack of consensus is evident in the disparate classifications across organizations. For example, 204 of the 480 unique entities are identified as carcinogens by only one of the above 5 organization^2–7^ (Supplementary Data 1 – spTable1.xlsx). This diversity in classification emphasizes the complexity of carcinogen identification and the need for a more streamlined, computational approach to handle this biomedical data deluge and ensure comprehensive public health protection.

There has been significant progress in this direction with the development of computational chemical informatics tools such as the RDKit^11^ and AI frameworks like Deepchem^12^ & ChemGPT^13,14^. However, streamlining these insights with conclusive literature evidence from scientific publications providing key contextual information regarding exposure, dosage, prevalent environmental and sociological factors, as well as mechanisms of action has yet to be achieved^3^. Additionally, there is a need for quantitation of non-chemical factors responsible for carcinogenicity^15^, which these tools are not designed to handle. For example, exposures found through epidemiological evidence, such as aflatoxins in processed wheat supply ^16–18^, Polychlorinated biphenyls (PCBs) in regional drinking water ^19–21^, chronic cancer associated with diseases like Hepatitis B & C^22–24^, and the effect of physiological stress through disruption of the sleep cycle^25–27^ are missed by such tools. Recent efforts have attempted to integrate text mining and database fusion approaches for prioritizing carcinogenic agents^28^. While these methods represent a step forward, they rely on relatively simple text mining techniques and database queries that may not fully capture the complex nuances of scientific literature. Moreover, their reliance on older natural language processing methods limits their ability to extract and synthesize information from the rapidly growing body of biomedical literature. There remains a need for more sophisticated methods that can leverage recent advances in Natural Language Processing (NLP), and machine learning to comprehensively analyze the available evidence.

NLP, particularly with the advent of transformer-based models, such as Large Language Models (LLMs) and ChatGPT, stand at the technological forefront, automating the distillation and categorization of pertinent information from plethora of text with remarkable precision^29–32^. Yet, these advanced models are also not impervious to the specialized jargon and complex nuances characteristic of scientific texts^33,34^, that still lead to inaccuracies and unintended biases^35,36^.

Addressing these limitations, we introduce the Carcinogen Detection via Transformers (CarD-T) framework, an innovative amalgamation of probabilistic statistical analysis^37^, and the contextual acumen of transformer-based learning in biomedical literature^15,38^. CarD-T leverages transformers for Named Entity Recognition (NER) tasks, achieving a balance of accuracy and computational efficiency that rivals extensive models like GPT-4, without imposing excessive computational demands. CarD-T is meticulously engineered to navigate and quantify the intricate narrative of scientific discourse, placing a spotlight on the task of nominating potential carcinogens. It seeks to capitalize on the strengths of transformer architecture, which is fine-tuned to harmonize performance with computational feasibility. This manuscript delves into the conception, development, and empirical validation of the CarD-T framework. By automating the recognition and nomination of carcinogenic entities, CarD-T aspires to streamline toxicogenomic literature reviews, providing a robust and scalable tool for the swift identification of potential carcinogens within the sprawling expanse of scientific literature.

## Approach

### CarD-T Framework: Schematic Overview

**Figure 1:**
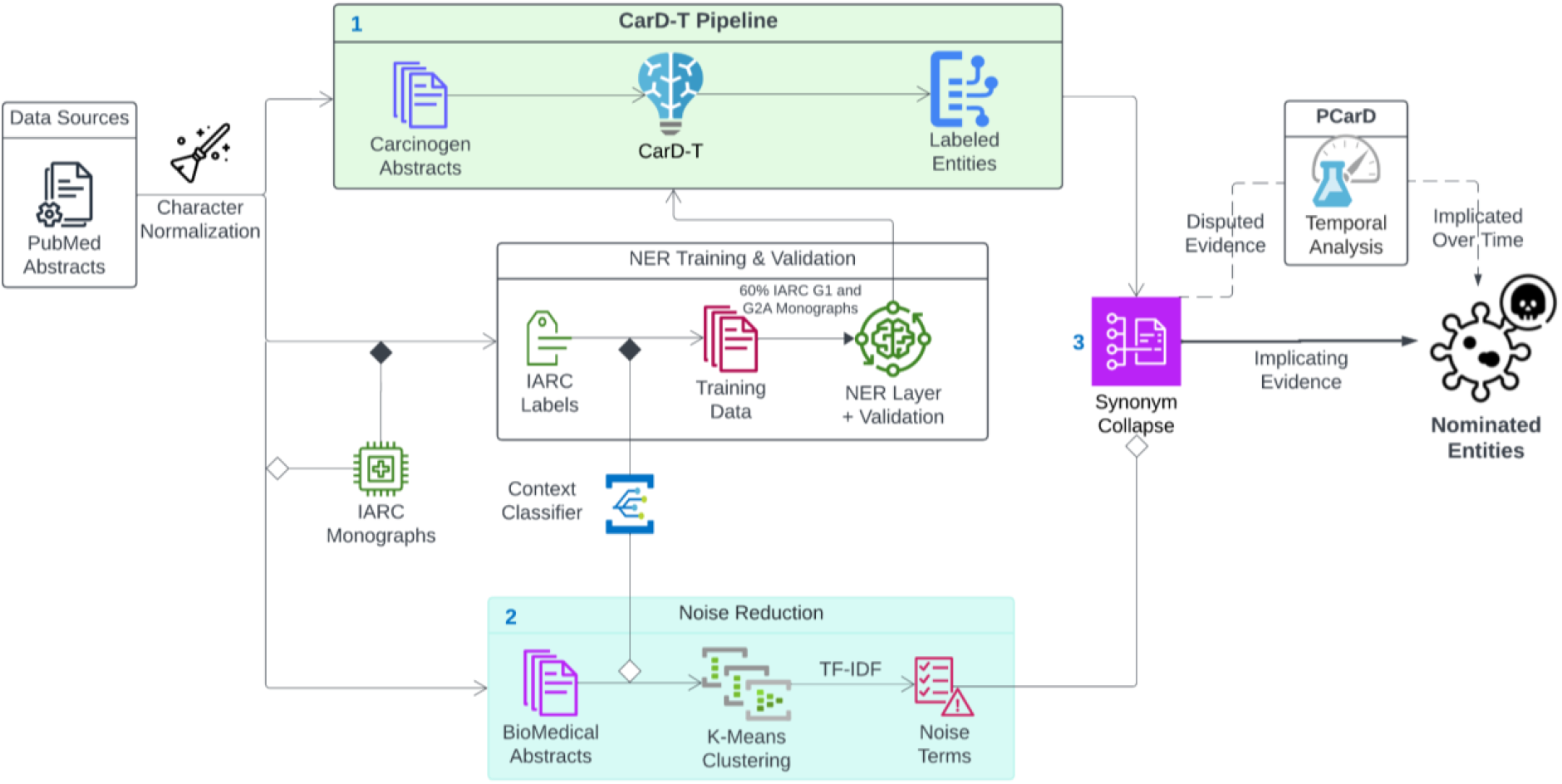
Schematic of the CarD-T Pipeline for the Identification and Nomination of Potential Carcinogens. The pipeline commences with data extraction from PubMed abstracts enriched with carcinogen-related keywords. At this point, there are two concurrent branches – 1) Named Entity Recognition Training and Validation – done on context derived from abstracts containing IARC G1 & G2A carcinogens, and 2) noise reduction using a Context-Derived TF-IDF approach abstracts clusters obtained via K-Means to refine the corpus. The NER model is trained and validated using labeled entities from the International Agency for Research on Cancer (IARC) Group 1 (G1) and Group 2A (G2A) carcinogens, with a context classifier integrated to enhance context-specific recognition of noise. The trained NER model is used to extract and label potential carcinogen entities from the broader carcinogen lexicon from the past 25 years. The noise terms identified by the noise reduction module are used to clean-up the labeled entities, providing a clean list of potential carcinogens to be assessed and analyzed, along with associated scientific publication records. Next, 3) Synonymous terms are collapsed into count data and collated over time. An additional temporal analysis of the associated publication data using the Probabilistic Carcinogen Denomination (PCarD) module, can provide consensus scores for each of the identifies carcinogen entities. The entire process is geared towards streamlining the identification of putative carcinogens from biomedical literature, offering a scalable and robust tool for contemporary toxicological studies.

### Data Sources and Training for Carcinogenic Entity Recognition

Our training data was derived from PubMed abstracts containing IARC monographs, as IARC has profound documentation, including all published sources, for the justification of their carcinogenic entities. Additionally, IARC stands out as the only major organization to include documentation regarding non-chemical carcinogenic exposures.

We began by collecting 236,696 abstracts published between 2000-2024 from PubMed with at least one citation, each indexed with top carcinogen-related Medical Subject Headings (MeSH, detailed in Table 1)^39,40^. To standardize our data, we employed normalization form compatibility decomposition to process special characters consistently^41^. This normalization included adjustments for journal-specific character variants and alignments with the International Union of Pure and Applied Chemistry (IUPAC) standards. For convenience of writing, we will refer to this data set in the following text as DS – Carcinogen.

**Table 1:** Overview of Carcinogen Nomination Contexts & their MeSH terms in Literature.

| Contextual Significance | Description | MeSH Terms |
| --- | --- | --- |
| Genotoxic | Involves damage to the genetic information within a cell causing mutations | DNA Damage, Mutagenesis, Genotoxicity |
| Mutagenic | Leads to genetic mutations | Mutagens, Genetic Mutations |
| Proangiogenic | Promotes the formation of new blood vessels | Neovascularization, Pathologic, Angiogenesis Inducing Agents, Cancer |
| Mitogenic | Stimulates cell division and growth | Cell Proliferation, Mitogens, Cancer |
| Alkylation | Introduces an alkyl group into molecules, potentially damaging DNA | Alkylating Agents, DNA Alkylation |
| Reactive Oxygen Species | Involves chemically reactive molecules containing oxygen | Reactive Oxygen Species, Oxidative Stress |
| DNA Adducts | Forms covalent bonds with DNA, leading to mutations | DNA Adducts, Carcinogens |
| Toxic or Oncogenic Organisms | Involves organisms that produce carcinogenic substances | Carcinogens, Biological, Oncogenic Viruses |
| Peroxisomal Proliferation | Increases the number of peroxisomes, affecting cellular metabolism | Peroxisome Proliferators, Cellular Metabolism |
| Oxidative Stress | Leads to cellular damage due to an imbalance between free radicals and antioxidants | Oxidative Stress, Antioxidants |
| Cancer Promoters | Substances that do not cause DNA mutations but promote the proliferation of cells with mutations, increasing the likelihood that cancer will develop. | Tumor Promoters, Carcinogenesis |
| Co-carcinogens | Substances that do not independently cause cancer but can act synergistically with other carcinogens to increase the risk of cancer. | Co-carcinogens, Carcinogenicity Tests |
| Procarcinogens | Precursors that become carcinogenic after metabolic conversion. | Procarcinogens, Carcinogens |
| Chronic Exposures and Behaviors | Long-term occupational or environmental exposures to carcinogenic factors | Occupational Exposure, Environmental Exposure, Carcinogens |

Next, we collated the abstracts with IARC Groups 1 (G1) and 2A (G2A) class carcinogens, noted for their definitive carcinogenicity evidence in humans (G1) and mammals (G2A) with probable human effects^3,4^. We also expanded our search for IARC entity labels leveraging all available synonyms, including known IUPAC variations, pharmaceutical brands, United Medical Language System (UMLS) terms, and various linguistic derivatives (Supplementary Data 2 – spTable2.xlsx). We used abstract text with these expanded labels list to subset potential training data from the total 236,696 carcinogen abstracts (referred to going forth as DS – G1,2A).

In addition, we also trained a few-shot learning based contextual classifier^42,43^ for segregating context, trained on Sentence-BERT^44^ (SBERT) embeddings from the Large General Text Embeddings^45^ (GTE) SBERT model. To achieve this, we manually identified 100 sentences from DS – G1, 2A that contained phrases showing clear “evidence of carcinogenicity” and tagged them as EC (see Table 1)^15,46^. Additionally, we identified 100 sentences from the DS-Carcinogen abstracts that showed clear “evidence of non-carcinogenicity” and another 100 sentences which, while mentioning a chemical entity, were ambiguous in its role in causing carcinogenesis from the DS-Carcinogen dataset. We labeled these contexts as ENC or UN (unknown) respectively. The few-shot learning classifier was trained on these 300 sentences and used to identify additional sentences with “evidence of carcinogenicity” or “evidence of non-carcinogenicity”. Using 100 sentences each from EC, ENC and UN control group, the classifier achieved an F1 score of 0.926. (Code available https://github.com/jimnoneill/CarD-Few based on the SetFit algorithm for contrastive “few-shot” learning, which emphasizes learning from the differences and similarities among examples.)

The few-shot learning-based classifier was used to then identify 60 EC sentences per G1 and G2A carcinogens from DS – G1, 2A. In cases where 60 EC sentences were not available, we had ChatGPT-4 generate additional synthesized EC sentences for these carcinogens modeled off our existing ones. (Code & prompts available https://github.com/jimnoneill/CarD-Gen). Additionally, the few-shot learning based classifier was used to identify 1000 ENC sentences from the DS – Carcinogen database. Once again, ChatGPT-4 was used to generate 9075 more synthetic ENC sentences with G1 and G2A carcinogens as subjects (55 ENC sentences for each carcinogen), based on the 1000 actual ENC sentences identified from the DS – Carcinogen text database. This was specifically done to prevent EC bias towards G1 and G2A in any future learning models. In all, we ended up with 19,975 sentences with G1 and G2A carcinogens primarily as subjects labeled as EC or ENC (approximately equal EC and ENC). The sentences that came from actual scientific abstracts were then expanded to contain the entire abstract. The sentences that were synthetic stayed as is. All of the text in this dataset was further processed manually to tag individual entities as “carcinogen-positive”, “carcinogen-negative”, “carcinogen-anti-neoplastic”, and “cancer-type”. This expanded data set is referred to as Context Training – DS going forth. 18,975 expanded sentences (full abstract) or synthetic sentences in this data set are each associated with one (or rarely two) G1 and G2A carcinogens. This data set is used for fine-tuning a Named-Entity-Recognition model to identify novel carcinogens continuously being reported on by the scientific community.

The selection of 60 EC and ENC texts per G1 and G2A IARC carcinogen is derived from methods previously described on ensuring balanced & sufficient representativeness of the entity types^47,48^. This number allows for capturing a variety of linguistic patterns and contexts, which is crucial for training robust NER models.

The total dataset size of 18,975 sentences is significant for training a high-performing NER model, especially one based on a transformer architecture like Bio-Electra, which requires extensive data to fine-tune^49,50^. Larger datasets can better capture the complexity and nuances of language, which is particularly important in the biomedical domain where precision is critical.

### Named Entity Recognition (NER) Training Methodology

To be able to annotate entities in scientific abstracts as EC or ENC, we trained and fine-tuned CarD-T, a 335 million parameter language model based on Bio-ELECTRA architecture^50^. Bio-ELECTRA stands out for its accuracy as an NER labeling transformer on the Biomedical Language Understanding & Reasoning Benchmarks (BLURB)^49^. As opposed to a generative transformer, it has features which are crucial for up-to-date quantitation of potential carcinogenic exposures. This architecture also offers lightweight, local deployment in less resource-intensive environments.

The training data for CarD-T was selected from the Context Training – DS by selecting text associated with a specific subset of G1 and G2A carcinogens. Using this data subset, CarD-T was trained to parse the input data and classify it as “carcinogen-positive”, “carcinogen-negative”, “carcinogen-anti-neoplastic”, and “cancer-type”. The training regimen for CarD-T included an early-stopping callback with a patience parameter set to 1 and a delta threshold of 0.01 to further prevent overfitting^51^. Optimization of the model’s weights was set with a maximum of 3 epochs using an AdamW optimizer^52^, with a carefully chosen learning rate of 2 × 10^−5 53^. The size of the training data subset from the Context Training – DS was iteratively selected from text associated with 20% of randomly selected G1 and 2A carcinogens to 90% of randomly selected G1 and 2A carcinogens, in increments of 5%. We found that CarD-T trained on data subset for 60% of G1 and 2A carcinogens was able to correctly identify 100% of G1 & G2A class carcinogens from the full Context Training - DS. Thus, we constrained the training set to 60% of the IARC G1 and 2A entities to account for the least over-fitting^54^. This selection process engendered a corpus comprising a total of 11,985 annotated text (including expanded sentences and synthetic sentences) for training. The remaining 40% of IARC G1 and 2A entities were consistently recognized in our validation set of 7,990 sentence corpus.

### The Carcinogen-Entity Nomination Extraction Process, including Noise Reduction

Next, we applied the CarD-T NER to annotate entities in the 216,721 remaining carcinogen abstracts from DS-Carcinogen. This process identified 11,129 unique entities that CarD-T classified as “carcinogen-positive”, were present in at least two separate PubMed articles, and excluded those already listed as known G1 & G2A entities (and their synonyms). Further entity processing involved lemmatization using SpaCy^55^, converting to lowercase, expanding abbreviations, and normalizing special characters and spaces. We limited the occurrence count of unique entities to one per abstract to prepare for Probabilistic Carcinogen Denomination (PCarD) classification of entities containing both “carcinogen-positive” and “carcinogen-negative” evidence across the abstracts^37^ (see details for PCarD below).

Due to the inherent limitations of NER, which can sometimes misclassify entity fragments^43,56^, we employed a de-noising step. For instance, NER might incorrectly separate “infected cells” from “HIV-1” in the phrase “HIV-1 infected cells shown increased patterns of genotoxicity”. To overcome this, we used Term Frequency-Inverse Document Frequency (TF-IDF)^57^ with a 4-word n-gram range to filter out irrelevant terms. When this proved inadequate, we advanced to a Context-Derived TF-IDF (CD-TF-IDF) method for more precise noise reduction^58^ as described below -

We obtained noise terms derived from a broader 1.5 million documents across biomedical fields, including oncology^39^. We segmented these into topics using K-means clustering on their SBERT embeddings, guided by the Calinski-Harabasz index for optimal cluster size^59^. In parallel, we used the few-shot trinary classifier to identify abstracts containing EC or ENC sentences. These abstracts were removed from the individual clusters they were grouped into. We excluded any clusters where more than 25% of the abstracts contained EC or ENC contexts. TF-IDF analysis was then conducted on each cluster to further differentiate noise terms. These noise terms were eliminated from the raw CarD-T – NER labeled entities. The CD-TF-IDF procedure eliminated 2,203 classified entities.

Among the remaining uniquely classified entities, we consolidated all synonyms with an *in house* algorithm, Synonym-Lustre, a tool for synonym disambiguation in biomedical terminology. This reduced our dataset 8926 labeled entities to 4321 entities. These contained 2,484 EC candidates and 2,391 ENC candidates, with an overlap of 554 entities tagged as both EC and ENC in different abstracts.

Details of these CarD-T identified EC and ENC entities, including all synonyms and PubMed Unique Identifiers (PMIDs), are compiled in Supplementary Data 3 – spTable3.xlsx.

The pathway to assess and analyze any past or future text to identify potential carcinogens indicated in scientific literature is summarized in figure 1.

**Figure 2:**
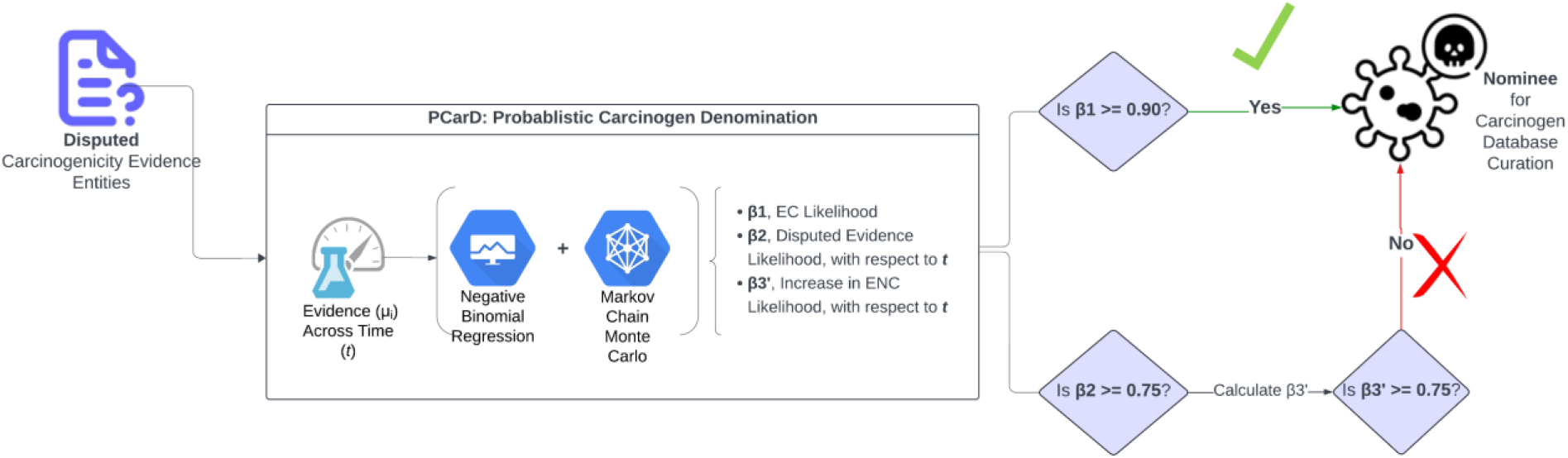
This flowchart illustrates the PCarD process for classifying ‘Disputed’ carcinogenic entities. It begins by fitting a negative binomial distribution to entity counts overtime and evaluating β0, β1, β2, β3 and β3′ from equation 1. If β1≥0.9, the entity is classified as ‘Nominee’ true, representing strong number of EC counts for that entity. However, if β2 ≥ 0.75, this suggests a strong temporal trend. At this point, β3 and β3′ are assessed. If β3′≥0.90, the entity is reclassified as ‘Disputed’ due to recent exculpating evidence; otherwise, it is maintained or updated to ‘Nominee’ based on the evidence.

### Evaluating CarD-T Framework’s Efficacy in Nominating New Carcinogens

The 2,484 EC nominees and 2,391 ENC nominees needed manual review for correct contextual classifications. To ameliorate the burden of manual vetting required, we employed a randomized sampling approach for finite populations^60^. We calculated our sampling process for sample size (n) as:

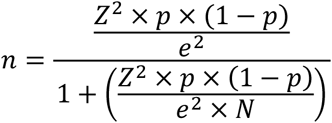

where *n* is the sample size, Z the confidence level Z-score, *p* the estimated proportion of the population, *e* the margin of error, and *N* the population size. We calculated a robust sample size for validation, where:

- Population Size: 2,484 unique EC entities; 2,391 ENC entities
- Sample Size: 290 EC; 288 ENC, derived using 95% confidence and a 5% margin of error
- Confidence Level: 95%
- Margin of Error: 5%

For the 290 & 288 randomly selected entities we tracked instances of True Positives (TP), False Positives (FP), and False Negatives (FN)^61^ to calculate precision, recall, and the harmonized F1 score as performance indicators.

### CarD-T vs GPT-4 on Annotating Carcinogens in-Context

We wanted to understand how well our framework performs compared to ChatGPT-4, in annotating carcinogens. We used a few-shot learning approach, with two examples per EC and ENC category, following similarly documented methods^62^ to mimic our annotation methods through ChatGPT-4. We manually evaluated 400 scientific abstracts, of which half abstracts were chosen for content related to EC or ENC, known to contain True Positive (TP) entities. The other half were selected to represent True Negative (TN) entities, intentionally excluding EC or ENC references. All abstracts were vetted to ensure the absence of G1 or G2A carcinogen classifications.

We conducted power analysis to determine an appropriate sample size for the comparison. This size ensures a reliable test for rejecting an incorrect null hypothesis, considering the given effect size and the level of statistical significance^63,64^.

The effect size was derived using the formula:

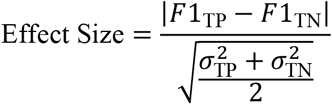

In this equation, *F*1_TP_ and *F*1_TN_ represent the aggregate F1 scores for the True Positive and True Negative abstract groups. The *σ*_TP_ and *σ*_TN_ denote the standard deviations of the F1 scores for these groups.

The power of our statistical test, aiming to avoid a Type II error, was calculated using a two-sample t-test with an alpha (α) level of 0.05^63^.

### PCarD: Probabilistic Carcinogenic Denomination for Disputed Entities

Next, for entities tagged as both EC and ENC, due to potential opposing evidence trends for carcinogenecity, going forth called as ‘Disputed’ entities, we wanted to provide probabilistic inference based on evidence trends. Thus, our total of 554 ‘Disputed’ carcinogenic entities were subject to Bayesian temporal Probabilistic Carcinogenic Denomination (PCarD) to assign likelihood for carcinogenic ‘Nominee’ status relative to changes in evidence sentiment over time. (Details ‘Disputed’ entities are cataloged in Supplementary Data 3 – spTable3.xlsx)

### Bayesian Model Specification

The observed counts of entity *Y*_*i*_, including both EC and ENC counts, as a function of elapsed time interval *t*_*i*_ was modeled using a Negative Binomial distribution to accommodate overdispersion observed in the data:

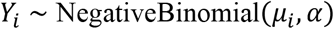

where:

- *μ*_*i*_ = *μt*_*i*_ represents the expected count for each entity after elapsed time, *t*_*i*_.
- *α* denotes the dispersion parameter of the distribution.

The expected count *μ*_*i*_ at time *t*_*i*_ is determined using Negative Binomial Regression as:

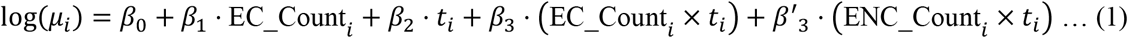

where:

- *t*_*i*_, is the relative elapsed time, centered around the mean year of the study period to address multicollinearity, reducing the correlation between the time variable and its interaction terms in the model.
- *β*_0_ is the intercept, representing the baseline log count.
- *β*_1_ quantifies the difference associated with existing carcinogen evidence.
- *β*_2_ captures the trend over elapsed time, *t*_*i*_, with each yearly interval centered around the mean to mitigate multicollinearity.
- *β*_3_ and *β*′_3_ assess how the interaction between the amount of evidence and the publication year affects the likelihood of an entity being classified as a carcinogen or not, respectively.

Priors for the regression coefficients were set as normally distributed, reflecting a lack of strong prior belief regarding their values:

- *β*_*k*_ for *k* = 0,1,2,3 are assumed to be normally distributed:

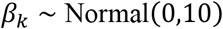
- The dispersion parameter *α* is assigned a Gamma prior to ensure positivity:

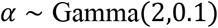

### Inference and Likelihood for Carcinogenic Denomination

To estimate each parameters’ likelihood probabilities, we employed a Markov Chain Monte Carlo (MCMC) technique. The MCMC method samples from the joint posterior distribution of the parameters by creating a chain of samples where each sample depends only on the previous one. Mathematically, the process can be represented as:

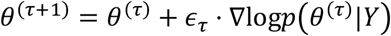

where *θ*^(*τ*)^ denotes the parameter values at step *τ, ∈*_*τ*_ is a small step size, and ∇log*p*(*θ*^(*τ*)^|*Y*) represents the gradient of the log posterior at *θ*^(*τ*)^. This iterative process refines the estimates of *β*_*k*_ and *α* parameters, ultimately allowing us to classify entities probabilistically as a ‘Nominee’ for carcinogen databases or not, based on the calculated posterior probabilities.

### PCarD: Nominee & Disputed Classifications

We used PCarD to re-classify ‘Disputed’ entities with a probability of *β*_1_ ≥ 0.90 as a ‘Nominee’ for carcinogen database curation. A *β*_2_ ≥ 0.75 signified a moderate-to-strong effect of time with changing evidence for ‘Disputed’ counts. If a moderate temporal effect was observed, *β*′_3_ value is considered. *β*′_3_ ≥ 0.90 likelihood is considered ‘Disputed’ regardless of high *β*_1_ likelihood, implying some recent rise of exculpating evidence of its carcinogenicity.

## Results

### Carcinogen Nominees: Navigating Our Supplementary Data of Carcinogenicity Evidence

**Supplementary Data 3 (spTable3.xlsx file)** accompanies this manuscript containing the following tables essential for assessing the identification and nomination of potential carcinogens as detailed in our study. Our data does not nominate entities from within our training data (any of IARC G1 and G2A class carcinogens).

1. **All Carcinogen nominees and associated Metadata (2000-2024, sheet 1 in the xlsx file)**: Table of 2,484 CarD-T labeled carcinogen entities and ‘Nominee’ and ‘Disputed’ classes based on evidence counts or PCarD classification if there was exculpating evidence. Each entity is listed with respective PMIDs for positive and negative conclusion counts, the years these studies were published, and additional contextual synonyms. (Synonym groupings by Synonym-Lustre were calibrated using IARC carcinogen synonyms, with 0.843 accuracy.)
2. **Bayesian Analysis (sheet 2 in the xlsx file)**: Analysis of ‘Disputed’ evidence trends from PCarD, where each entity is evaluated for its likelihood of being a carcinogen relative to evidence discourse over time. The results include likelihood probabilities representing their current trends in literary discourse.
3. **Bayesian Metadata (sheet 3 in the xlsx file)**: Meta-analysis of Bayesian computations, detailing the model parameters, data inputs, and the statistical contexts used for analysis.

Researchers are encouraged to utilize these data to glean insight & understanding of the dynamics influencing carcinogen research over the previous 25 years through practical application of this dataset in both ongoing and future studies.

### CarD-T NER is Calibrated on G1 & G2A Class Carcinogens

As CarD-T NER training data increased from 20% to 60%, the model showed significant improvements across all metrics, achieving a 100% success rate in identifying IARC class carcinogens at 60% data usage (Table 2).

**Table 2:** Cross-validation results for different increments with linear adjustments and random fluctuations, including the percentage of IARC Monographs identified.

| Increment | Accuracy | Precision | Recall | F1 Score | IARC Found (%) |
| --- | --- | --- | --- | --- | --- |
| 20% | 0.630 | 0.568 | 0.586 | 0.579 | 51.2 |
| 40% | 0.783 | 0.714 | 0.734 | 0.725 | 87.8 |
| 60% | 0.968 | 0.870 | 0.909 | 0.889 | 100.0 |
| 80% | 0.972 | 0.887 | 0.929 | 0.908 | 100.0 |
| 100% | 0.994 | 0.873 | 0.915 | 0.894 | 100.0 |

Beyond the 60% data mark, the model’s performance remains stable, with minor fluctuations in precision but steady gains in accuracy and recall, consistently identifying 100% of IARC G1 & G2A classes (Table 2). This suggests that additional data beyond this point does not markedly enhance the model’s recognition capacity, indicating an optimal data usage threshold at 60%.

In essence, the model’s evolving performance and consistent identification of IARC G1 and G2A underscore the importance of adequate data utilization. The CarD-T model excels in generalizing from training data to accurately detect carcinogenic entities, especially evident when utilizing 60% of training data, highlighting the significance of data volume and quality for optimal model performance.

### CarD-T Framework Successfully Identifies Nominees in-Context

The CarD-T NER yielded 2,484 & 2,391 EC & ENC (implicating & exculpating evidence) entities with 554 overlapping entities. Each of the entities are linked to a several unique PMIDs associated with their contextual mentions (EC nominees listed in Supplementary Data 3, ENC nominees not listed as they are not relevant here).

The CarD-T framework’s ability to identify novel carcinogens currently not in major databases was subject to manual verification, yielding F1, precision, and recall scores of 0.823, 0.793, and 0.857, respectively, for EC entity counts. (Figure 3, Table 3).

**Table 3:**
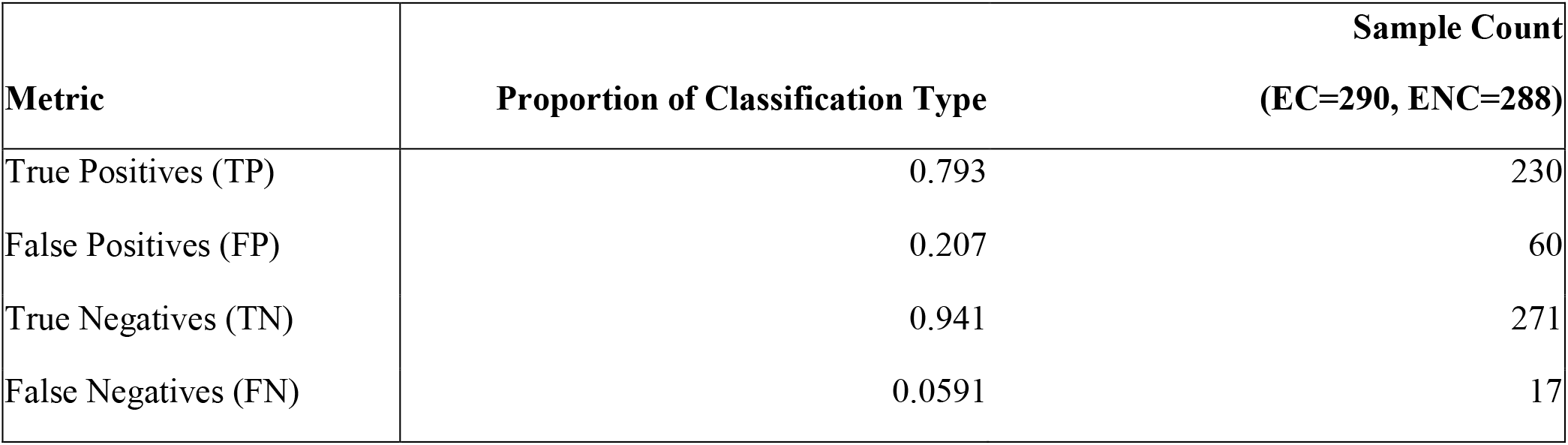
Classifications by proportion of TP & FP of EC & ENC entities from manually vetted cancer & carcinogenesis abstracts from 2000-2024 with 95% confidence and 5% margin of error – shown for one out of five samples. EC entities are Positives and ENC entities are Negatives.

**Figure 3:**
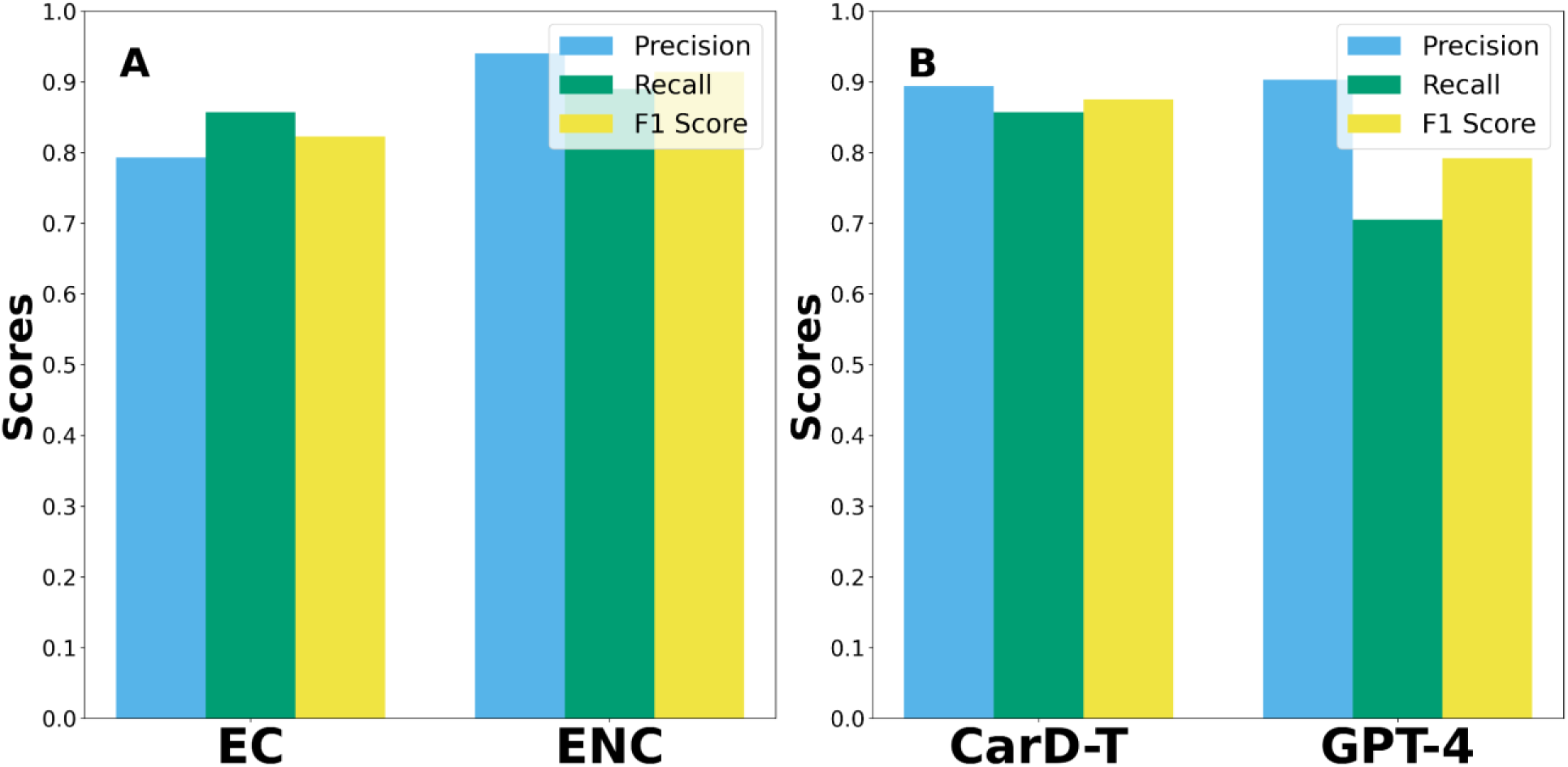
A) This bar graph contrasts the precision, recall, and F1 scores of context validation for contextually finding and refuting novel carcinogens not existing in current databases. Interestingly, metrics for EC contexts are significantly lower than ENC with F1, precision, and recall scores of 0.822, 0.789, and 0.857 versus 0.914, 0.940 and 0.890, respectively. B) This bar graph contrasts the precision, recall, and F1 scores of CarD-T and GPT-4 in identifying EC and ENC entities from any input scientific abstracts. While GPT-4 shows a marginally higher precision, CarD-T excels in recall. The harmonized F1 scores further reveal CarD-T’s superior performance in accurately identifying carcinogens.

Our aim was to identify & nominate potential carcinogens in literature not curated by existing IARC monograph evidence or belonging to our training data. Incidentally, False Positives (FP) represented one fifth of our results for EC entities. Out of these roughly one third were variants of G1 & G2A our noise reduction methods failed to recognize. Roughly 45% of FP were hypernymy descriptions of entities consisting of broad terms like ‘organic extract’ and ‘soluble particle’. The remaining FP were incorrectly classified entities.

ENC context counts yielded more impressive validation of 0.914, 0.940 and 0.890 for F1, precision and recall, respectively (Figure 3, Table 3A).

### CarD-T Harmonically Outperforms GPT-4

A comparative study assessed CarD-T’s and GPT-4’s performance on a subset of 400 abstracts, that specifically excluded any abstracts associated with G1 and G2A IARC carcinogens or their synonyms, for labeling potential carcinogen nominees. CarD-T’s precision was nearly on par with GPT-4’s, with scores of 0.894 and 0.903, respectively. However, CarD-T demonstrated superior recall at 0.857, compared to GPT-4’s 0.705, indicating more effective annotation of contextually relevant carcinogens. This precision-recall dynamic is further elucidated in Figure 4. The harmonized F1 scores stood at 0.875 for CarD-T and 0.792 for GPT-4, underscoring CarD-T’s proficient performance (Figure 3B).

### PCarD Analysis Identifies Temporal Trends in Carcinogen Research

554 entities had count types for both EC & ENC linked to multiple PMIDs and were categorized as ‘Disputed’ (Supplementary Data 3).

After comparing evidence trends with respect to time, PCarD re-classified 76 of these ‘Disputed’ entities into ‘Nominees’ to be considered alongside other only EC entities.

Each entity’s classification as Nominee or Disputed is accompanied by counts of publications affirming or challenging their carcinogenicity, with EC & ENC representing counts of evidence from 2000-2024, respectively (Table 4; Supplementary Data 3). Colors in Table 4 are to place emphasis on various trends. (Yellow—implicating; Red—exculpating; Purple—inconclusive.)

**Table 4:** Sample of Selected Disputed Nominees and Trends.

| Disputed | $\beta_1$ | $\beta_2$ | $\beta_3$ | $\beta_3'$ | EC Count | ENC Count | Nominated |
| --- | --- | --- | --- | --- | --- | --- | --- |
| singlet oxygen | 1.0 | 0.920 | 0.0108 | 0.993 | 38 | 2 | FALSE |
| schizophrenia | 1.0 | 0.742 | 0.733 | 0.262 | 49 | 2 | TRUE |
| curcumin | 0.0 | 1.0 | 0.930 | 0.0692 | 124 | 1702 | FALSE |
| bleomycin | 0.525 | 0.359 | 0.0 | 0.0683 | 211 | 211 | FALSE |
| mir 1290 | 0.0547 | 0.789 | 0.840 | 0.176 | 12 | 22 | FALSE |

#### Sample Assessment of Selected Disputed Entities from Table 4

##### Singlet Oxygen

Singlet oxygen shows a unique trend where it has a strong affirmative evidence count (β1=1.0) and a notable time-related trend (β2=0.920). However, the interaction effect (β3′=0.993) reveals a significant recent rise in exculpating evidence, leading to its classification as ‘Disputed’. This trend could indicate recent shifts in research focus or new findings that challenge previously held beliefs about its carcinogenicity, making singlet oxygen an interesting case of evolving scientific consensus.

##### Schizophrenia

Schizophrenia presents a high affirmative evidence count (β1=1.0) and a strong temporal trend (β2=0.742). The significant β3 value (0.733) suggests consistent supporting evidence over time.

This classification as ‘Nominee’ may reflect growing recognition of the potential carcinogenic links associated with schizophrenia, possibly through genetic or environmental factors that warrant further investigation. The relatively lower β3′ (0.262) indicates less recent negative evidence, reinforcing its classification as ‘Nominee’.

##### Curcumin

Curcumin stands out with its high temporal trend (β2=1.0) but remains classified as ‘Disputed’ due to an overwhelming amount of exculpating evidence (ENC Count = 1702) compared to supporting evidence (EC Count = 124). The high β3 (0.930) indicates substantial supporting evidence, but the significant volume of negative evidence underscores the conflicting nature of research on curcumin’s carcinogenicity. This duality makes curcumin a compelling subject for further study, as it exemplifies the complexity of interpreting mixed scientific evidence.

##### Bleomycin

Bleomycin exhibits moderate affirmative evidence (β1=0.525) and lower temporal trends (β2=0.359). The equal counts of affirmative and negative evidence (211 each) highlight its contentious nature in carcinogenic studies. The negligible interaction effect (β3′=0.0683) further indicates a lack of significant recent shifts in evidence, suggesting that bleomycin’s carcinogenicity remains an open question. This balance of evidence makes it a notable example of a chemical with long-standing scientific debate.

##### mir 1290

mir 1290, a microRNA gene and potential oncogene, has low affirmative evidence (β1=0.0547) but shows a moderate temporal trend (β2=0.789). The strong β3 (0.840) indicates a significant amount of supporting evidence over time. However, the moderate negative interaction effect (β3′=0.176) suggests some recent challenges to its carcinogenic classification. With relatively low EC and ENC counts (12 and 22, respectively), mir 1290 represents an emerging area of research, where ongoing studies may provide more clarity in the future.

## Discussion

The CarD-T framework, complemented by our Probabilistic Carcinogen Denomination (PCarD) module, has demonstrated exceptional prowess in identifying potential carcinogens within a vast corpus of scientific literature. Our analysis yielded approximately 1930 pure EC (evidence of carcinogenicity) entities, not including the 554 disputed cases. While we estimate that about 20% of these are false positives due to various factors discussed earlier, this still leaves us with close to 1500 potential carcinogens worthy of further investigation.

Examining the list of the CarD-T EC (Evidence of Carcinogenicity) nominees reveals a diverse array of entities, spanning from drugs and biochemical compounds to social, behavioral, and environmental factors. For instance, our analysis flagged entities such as “polycystic ovary syndrome,” “non-alcoholic fatty liver disease,” and “micro plastics” alongside more traditional chemical compounds. This diversity highlights the complex, multifaceted nature of carcinogenesis and the importance of considering a wide range of potential contributing factors. Many of these nominees are not currently listed in major carcinogen databases, suggesting that they may represent emerging areas of concern or contentious cases requiring further study. For example, “ochratoxin A,” a mycotoxin found in various food products, was identified in our analysis with 162 supporting publications spanning from 2000 to 2022. Similarly, “borna disease virus” emerged as a nominee with 144 supporting publications from 2000 to 2022. Overall, an estimated 1476 entities identified as EC by the CarD-T algorithm, are not currently included in the list of entities classified with certainty to be carcinogens by any agency. These are after removal of 54 NTP, EPA, ECHA and OSHA (supplementary data 1) entities. Out of these remaining 1476 EC entities, 99 entities are those listed by IARC as Group 2B or Group 3 classification. It needs to be noted that all of these potential carcinogen nominees are described as such in at least 2 peer-reviewed and cited manuscripts. While, by some measures, this might still be a large list, the automated assessment of a corpus of over 200,000 scientific abstracts to obtain a compact list of less than 2000 potential carcinogenic factors, their possible synonyms and hyponyms, and references to associated manuscript identifiers, in our opinion, is a significant achievement. The fact that the involved computational pipeline can be deployed locally on a desktop GPU based computer with 64 GB of RAM (recommended), with approximately six hours of compute time (including the initial data sourcing from PubMed), makes CarD-T an extremely useful tool for any cancer, health and public policy researcher.

Interestingly, one of the top nominated EC entities is COVID-19, which warrants careful interpretation. While this entity was classified as carcinogenic with strong certainty based on contextual analysis, it’s crucial to note that the number of supporting publications (255 from 2020 to 2024) was significantly influenced by the unprecedented surge in research related to the pandemic during our study period. This underscores the importance of considering not just the quantity but also the timing and context of research outputs in our analysis.

The presented approach has its limitations. Foremost, the entities identified by CarD-T are potential candidates for carcinogenicity, not confirmed carcinogens. Additional processing, either through manual review or more sophisticated AI-based tools, is necessary to validate these findings. Furthermore, our current analysis is limited to abstracts, which may not capture the full nuances of study methods, result quality, or contextual details present in full-text articles.

Another limitation is the potential for bias introduced by publication trends, as illustrated by the COVID-19 example. Sudden surges in research interest around particular topics could lead to their overrepresentation in our results. Additionally, while our tool is effective at identifying potential carcinogens, it does not inherently assess the strength or quality of the evidence supporting each nomination.

Despite these limitations, our work opens up exciting opportunities for future research. We have envisioned and partially implemented sub-labels for the “cancer type” label in CarD-T NER, including “mechanism of action,” “study model (human vs. animal vs. population),” and “organ affected.” While the outcomes from these more granular classifications are still works in progress, they represent a promising direction for enhancing the tool’s capabilities. These sub-classifications could provide invaluable insights into the specific pathways and contexts in which potential carcinogens operate. For instance, they could help differentiate between agents that primarily affect certain organs or those that operate through particular cellular mechanisms. This level of detail could greatly assist researchers in prioritizing further investigations and potentially inform more targeted prevention strategies.

## Conclusions

We present CarD-T, augmented by PCarD, as an automated text analytics tool to assess and tabularize evolving evidence for and against known, suspected and unknown carcinogens from scientific corpus. The tool is an amalgamation of various existing and newly developed code that can be downloaded for free from our open-access repositories and deployed locally on any standard GPU based computer. The comparative analysis of this tool with GPT-4 has highlighted the distinct capabilities of CarD-T in the domain of carcinogenic entity identification from an ever-growing corpus on cancer incidence and associated causal factors. The model’s nuanced understanding of the scientific lexicon distinguishes true carcinogen research from incidental mentions, emphasizing its critical role in public health informatics. Thus, we believe that CarD-T can become an essential tool for regulatory agencies tasked with assessing and establishing the cancer-causing potential of various biochemical, environmental, social and occupational factors. At the same time, it can be deployed by individuals wishing to delve into carcinogenesis research and look at relationships between and across various factors associated with increased cancer risk and incidence.

Despite its impressive performance, CarD-T’s limitations point to areas for future improvement. The challenges posed by the subtleties and complexities of scientific literature necessitate continuous model updates and refinements. The integration of diverse data sources and multilingual datasets could further enhance the framework’s utility, providing a more comprehensive global assessment of carcinogenic agents.

Our tool’s unique niche in the public health informatics ecosystem complements the broader Biomedical NLP services offered, focusing specifically on the detection and tracking of research evidence implicating carcinogens across time. The insights gleaned from the CarD-T framework, particularly when augmented by Probabilistic Carcinogenic Denomination (PCarD), reflect the dynamic and evolving nature of carcinogenicity research, offering a valuable perspective for agile and informed public health responses.

## Data Availability

The datasets generated and/or analyzed during the current study are available from the corresponding author on reasonable request.

https://huggingface.co/jimnoneill/CarD-T

## Code & Data Availability

Nominated carcinogens from carcinomic lexicon spanning the last 25-years and their metadata are available in the supplementary data. The code for the CarD-T framework & PCarD, including the training and evaluation scripts, is available upon request from the corresponding author. Further details and access instructions will be provided in the final published version of this paper. CarD-T language model is available at https://huggingface.co/jimnoneill/CarD-T.

## Declarations

### Ethics Approval and Consent to Participate

Not applicable.

### Consent for Publication

Not applicable.

### Authors’ Contributions

J.O, A.G.R., N.D, L.A and P.K contributed to the conception and design of the study. J.O developed the CarD-T pipeline. J.O. and O.T. developed PCarD. J.O. performed the data collection and analysis. J.O, O.T and P.K. contributed to the writing of the manuscript. All authors read and approved the final manuscript.

## Acknowledgements

P.K. would like to thank Mr. Prasad Kothari for an introduction to NLP and text analytics and stimulating conversations on related topics. J.O would like to acknowledge support from the SDSU ACCEL program. P.K. and J.O. would like to acknowledge support from the SDSU HealthLINK center.

